# PhenoScore: AI-based phenomics to quantify rare disease and genetic variation

**DOI:** 10.1101/2022.10.24.22281480

**Authors:** Alexander J M Dingemans, Max Hinne, Kim M G Truijen, Lia Goltstein, Jeroen van Reeuwijk, Nicole de Leeuw, Janneke Schuurs-Hoeijmakers, Rolph Pfundt, Illja J Diets, Joery den Hoed, Elke de Boer, Jet Coenen-van der Spek, Sandra Jansen, Bregje W van Bon, Noraly Jonis, Charlotte Ockeloen, Anneke T Vulto-van Silfhout, Tjitske Kleefstra, David A Koolen, Hilde Van Esch, Gholson J Lyon, Fowzan S Alkuraya, Anita Rauch, Ronit Marom, Diana Baralle, Pleuntje J van der Sluijs, Gijs W E Santen, R Frank Kooy, Marcel A J van Gerven, Lisenka E L M Vissers, Bert B A de Vries

**Author notes:** corresponding authors (both authors contributed equally).

## Abstract

While both molecular and phenotypic data are essential when interpreting genetic variants, prediction scores (CADD, PolyPhen, and SIFT) have focused on molecular details to evaluate pathogenicity — omitting phenotypic features. To unlock the full potential of phenotypic data, we developed PhenoScore: an open source, artificial intelligence-based phenomics framework. PhenoScore combines facial recognition technology with Human Phenotype Ontology (HPO) data analysis to quantify phenotypic similarity at both the level of individual patients as well as of cohorts. We prove PhenoScore’s ability to recognize distinct phenotypic entities by establishing recognizable phenotypes for 25 out of 26 investigated genetic syndromes against clinical features observed in individuals with other neurodevelopmental disorders. Moreover, PhenoScore was able to provide objective clinical evidence for two distinct *ADNP*-related phenotypes, that had already been established functionally, but not yet phenotypically. Hence, PhenoScore will not only be of use to unbiasedly quantify phenotypes to assist genomic variant interpretation at the individual level, such as for reclassifying variants of unknown clinical significance, but is also of importance for detailed genotype-phenotype studies.

## 1 Introduction

A significant portion of individuals with clinically and genetically heterogeneous rare diseases, such as neurodevelopmental disorders (NDD), has been molecularly diagnosed in the last decade using whole-exome sequencing (WES) [1–4]. Clinical WES data interpretation relies on filtering and prioritization for rare genetic variants in disease-gene panels, which are subsequently interpreted in the context of the patient’s clinical presentation [5]. Whereas this strategy is essential to identify the disease-causing variant(s), it is estimated that, depending on the number of genes included in the panel, dozens of variants are prioritized as diagnostic noise [6] — and this number is expected to rise even more in the coming years with technological innovations such as genome sequencing finding their way into the diagnostic arena [7–9].

At the molecular level, several computational methods, such as MutationTaster [10], PolyPhen [11], SIFT [12], CADD score [13], have been designed to predict variant pathogenicity. These tools use diverse approaches, such as looking at the impact of the variant on protein structure (MutationTaster, PolyPhen), taking conservation into account (MutationTaster, PolyPhen, SIFT) — or trying to incorporate multiple sources of genomic information (CADD score). At the phenotypic level, headway has been made by introducing Human Phenotype Ontology (HPO), systematically capturing the presence of features observed in individuals with rare diseases [14]. However, equivalent to molecular tools, algorithms using these HPO data to quantify phenotypic HPO similarity between individuals with genetic disorders would provide significant benefits to diagnose rare disease. Such a quantitative phenotypic score could for instance assist with the interpretation of genetic variants of unknown clinical significance (VUS), which constitute 10-30% of all variants clinically assessed [4, 15]. Reducing the number of VUSs is of essence since studies have shown that not all individuals and families respond similarly to the result of a VUS test-result, and usually do not fully comprehend its meaning [16, 17], potentially leading to frustration, and/or distress due to the uncertainty involving a possible diagnosis and course of disease. Importantly, VUSs have also been shown to inflict inappropriate medical decisions [18, 19].

Next to reclassifying VUSs, quantifying phenotypic HPO similarity at the cohort level could also help to provide further steps towards personalized medicine by automatically recognizing distinct phenotypic subtypes leading to more tailored clinical prognosis [20–22].

A branch of science that could assist in objectively quantifying phenotypic data is artificial intelligence (AI). AI has dramatically reformed the manner clinical data are processed and analyzed in recent years, with the AI revolution in medicine starting in pathology and radiology [23–26]. In genetics, these new techniques have been employed in assisted interpretation of genomic variants [27–29] and combining molecular and phenotypic evaluations, mainly looking at methods to use phenotypic data in HPO to automatically prioritize genetic variants [30–36]. Furthermore, advances in computer vision have led to the application of facial recognition technology in clinical genetics [37–42]. Facial recognition is able to assist in the recognition of (neuro)developmental syndromes, since the development of the brain and facial shape are closely linked [43–46] — and therefore, it comes as no surprise that a significant part of genetic disorders have distinct facial features [47]. However, not all genetic syndromes have a clear, recognizable facial gestalt, which hinders methods solely looking at facial features. Moreover, a syndromic phenotype often includes more than ‘just the face’. Whereas tools have previously looked at either combining molecular data with either HPO, or alternatively, with facial features [1, 39], an important area has been left unexplored, which combines the facial- and HPO data into an AI-framework to predict phenotypic similarities without the need for genomic data input. Therefore, we developed PhenoScore: a next-generation open-source phenomics framework combining facial recognition technology with clinical features, quantitatively collected in Human Phenotype Ontology (HPO) from deep phenotyping.

## 2 Results

### 2.1 The PhenoScore framework

PhenoScore is a framework that currently consists of two modules: a component that extracts the facial features from a 2D facial photograph and a second module that takes HPO-based phenotypic similarity into account (Figure 1). The AI-based framework joins these results in three outputs: a Brier score and corresponding *p*-value, defining the individual’s clinical similarity to the syndrome assessed; a facial heatmap, highlighting important facial features for the syndrome; and, a visualization of the most important other (non-facial) clinical features. In the training phase of PhenoScore, at first an age-, sex-, ethnicity-matched dysmorphic control is sampled from our in-house database for every individual with the genetic syndrome of interest. Next, the facial features are automatically extracted from the facial photographs for both affected individuals and controls and the phenotypic HPO similarity is calculated (with several HPO terms and their child terms first removed from the dataset, as these are either facial HPO terms to be processed by the facial recognition module, or HPO terms that are deemed subjective and therefore at risk for interobserver variability). A support vector machine (SVM), a widely used classification algorithm in machine learning, is trained on these features, resulting in a trained classifier that can be used to generate a score for individuals, suspected to have the syndrome of interest. If we are interested in quantifying phenotypic (sub)groups, a permutation test is added during the training phase, determining whether the trained classifier performs better than random chance — providing evidence whether the two groups are distinguishable by PhenoScore. Finally, to provide insight into what PhenoScore is doing and to learn more about the investigated syndromes, explainable AI is incorporated into PhenoScore as well, using Local Interpretable Model-agnostic Explanations (LIME) [48, 49]. LIME works by generating random perturbed input data and inspecting the change in predictions, thereby obtaining data on the relative importance of each feature. By using LIME for both the facial- and HPO data, PhenoScore can generate facial heatmaps and visualizations on the most important clinical features.

**Figure 1:**
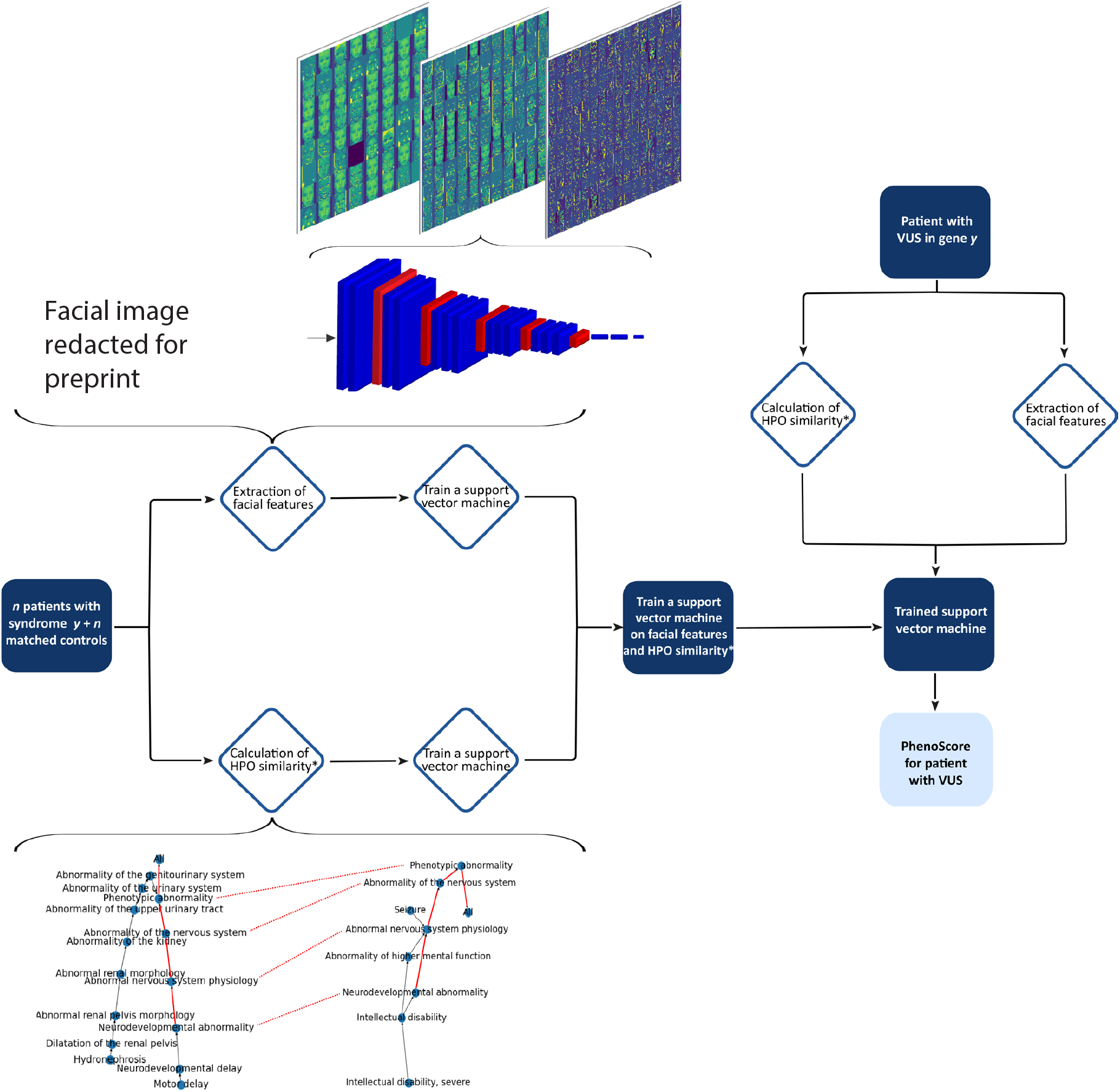
Here, the global workflow of this study is displayed, with the training and construction of PhenoScore on the left side. *n* individuals and *n* age-, sex- and ethnicity matched controls are selected for each syndrome. The facial features are extracted using a convolutional neural network, VGGFace2, and a support vector machine (SVM) is trained on these features. In parallel, the phenotypic similarity of individuals and controls is calculated, and a SVM is trained on those scores. Finally, a SVM is trained on both the facial features and the HPO similarity combined. On the right side of the figure, the trained classifier is used for a new individual with a VUS. Again, the phenotypic similarity and facial distances are calculated, and these are used as input for the trained SVM. The output is a score and assesses whether the individual of interest has that specific syndrome, thus the VUS being (likely) pathogenic.

### 2.2 Proof-of-Concept using PhenoScore for Koolen-de Vries syndrome

First, we investigated whether using our combined PhenoScore was actually an improvement on solely using either facial- or phenotypic data. The SVM was trained on both separate feature sets alone (e.g. HPO and facial features) and subsequently compared with the classification performance of PhenoScore. To measure classification performance, the Brier score [50] was chosen as the performance measure to focus on: it is defined as the mean squared difference between predicted outcome and observed actual outcome (lower is better). Next to that, we also report the area under the receiving operator curve (AUC; higher is better).

To demonstrate the power of the PhenoScore framework, we first performed a proof-of-concept study using 63 individuals with Koolen-de Vries syndrome (KdVS, OMIM #610443, Figure 2), caused by either proven pathogenic loss-of-function variants in *KANSL1* (*n*=11) or the 17q21.31 microdeletion (*n*=52). KdVS most prominent features reported in literature include hypotonia, intellectual disability, and joint laxity [51–53], for which the interdependence in our modelling is preserved using the graph structure of the HPO terms (Figure 2). Running Phenoscore on the 63 individuals with KdVS, we confirm the improvement on overall predictive performance when using both facial and clinical features compared to using either one alone (Brier score 0.106 or AUC 0.92 for PhenoScore, in contrast to 0.130/0.90 when using only facial data and 0.121/0.90 when using only phenotypic data, Table 1).

**Table 1:** The number of individuals per genetic syndrome included in our analysis are shown here. For every individual, a facial photograph, phenotypic data, and an age-, sex- and ethnicity control with a neurodevelopmental disorder is available (otherwise, the individual was excluded). Per genetic syndrome, the sex distribution, the median age and the results of the support vector machine (SVM) classifier are displayed here. The Brier score, for which lower is better, per syndrome is shown — with the numbers shown corresponding to the mean of the scores during the ten iterations in which matched controls were sampled. For almost all syndromes, the combination of facial- and phenotypic data is an improvement over using either dataset alone. Furthermore, the last column of this table displays the calculated *p*-values for the investigated syndromes using the random permutation test. All but one are significant at the 0.05 (and 0.01) level, as expected when inspecting the classification results.

| Gene/genetic syndrome | OMIM number | Number of individuals | Male/female (%) | Age (median in years) | Facial data only | HPO data only | PhenoScore | $p$-value |
| --- | --- | --- | --- | --- | --- | --- | --- | --- |
| 22q11 deletion syndrome | 611867 | 19 | 10/9 (53%/47%) | 5.0 | 0.189 | 0.146 | 0.124 | <0.001 |
| ACTL6A | NA | 3 | 2/1 (67%/33%) | 6.0 | 0.255 | 0.299 | 0.291 | 0.98 |
| ADAT3 (NEDBGF) | 615286 | 6 | 3/3 (50%/50%) | 7.5 | 0.180 | 0.075 | 0.059 | <0.001 |
| ADNP (Helsmoortel-van der Aa syndrome) | 615873 | 33 | 15/18 (45%/55%) | 5.0 | 0.209 | 0.156 | 0.145 | <0.001 |
| ANKRD11 (KBG syndrome) | 148050 | 22 | 15/7 (68%/32%) | 9.5 | 0.236 | 0.196 | 0.180 | <0.001 |
| ARID1B (Coffin-Siris syndrome) | 135900 | 34 | 16/18 (47%/53%) | 6.0 | 0.155 | 0.087 | 0.070 | <0.001 |
| CHD3 (Snijders Blok-Campeau syndrome) | 618205 | 27 | 11/16 (41%/59%) | 10.0 | 0.216 | 0.148 | 0.144 | <0.001 |
| CHD8 (IDDAM) | 615032 | 20 | 15/5 (75%/25%) | 11.0 | 0.254 | 0.174 | 0.163 | <0.001 |
| DDX3X (MRXSSB) | 300958 | 29 | 0/29 (0%/100%) | 8.0 | 0.171 | 0.030 | 0.025 | <0.001 |
| DYRK1A (MRD7) | 614104 | 13 | 7/6 (54%/46%) | 12.0 | 0.258 | 0.192 | 0.174 | <0.001 |
| EHMT1 (Kleefstra syndrome) | 610253 | 29 | 12/17 (41%/59%) | 6.0 | 0.220 | 0.107 | 0.090 | <0.001 |
| FBXO11 (IDDFBA) | 618089 | 18 | 14/4 (78%/22%) | 7.0 | 0.265 | 0.250 | 0.247 | <0.001 |
| KANSL1 (Koolen-De Vries syndrome) | 610443 | 63 | 28/35 (44%/56%) | 6.0 | 0.130 | 0.121 | 0.106 | <0.001 |
| KDM3B (Diets-Jongmans syndrome) | 618846 | 13 | 7/6 (54%/46%) | 7.0 | 0.262 | 0.185 | 0.194 | <0.001 |
| MECP2 duplication (MRXSL) | 300260 | 5 | 5/0 (100%/0%) | 8.0 | 0.169 | 0.214 | 0.173 | <0.001 |
| MED13L (MRFACD) | 616789 | 22 | 13/9 (59%/41%) | 6.0 | 0.235 | 0.140 | 0.124 | <0.001 |
| PACS1 (Schuurs-Hoeijmakers syndrome) | 615009 | 15 | 10/5 (67%/33%) | 4.0 | 0.240 | 0.146 | 0.140 | <0.001 |
| PHIP (Chung-Jansen syndrome) | 617991 | 16 | 9/7 (56%/44%) | 12.0 | 0.236 | 0.274 | 0.242 | <0.001 |
| PPM1D (Jansen-de Vries syndrome) | 617450 | 11 | 5/6 (45%/55%) | 7.0 | 0.269 | 0.189 | 0.163 | <0.001 |
| PURA (NEDRIHF) | 616158 | 33 | 18/15 (55%/45%) | 9.0 | 0.232 | 0.126 | 0.110 | <0.001 |
| SATB1 (truncating, DEFDA) | 619228 | 8 | 3/5 (38%/62%) | 6.5 | 0.260 | 0.170 | 0.164 | <0.001 |
| SATB1 (missense, Kohlschütter-Tonz syndrome-like) | 619229 | 11 | 5/6 (45%/55%) | 11.0 | 0.123 | 0.231 | 0.229 | <0.001 |
| SON (ZTTK syndrome) | 617140 | 25 | 13/12 (52%/48%) | 6.0 | 0.234 | 0.133 | 0.123 | <0.001 |
| TRIO (MRD63) | 618825 | 8 | 3/5 (38%/62%) | 10.5 | 0.161 | 0.157 | 0.159 | <0.001 |
| WAC (DeSanto-Shinawi syndrome) | 616708 | 9 | 3/6 (33%/67%) | 4.0 | 0.185 | 0.204 | 0.176 | <0.001 |
| YY1 (Gabriele-de Vries syndrome) | 617557 | 9 | 5/4 (56%/44%) | 9.0 | 0.175 | 0.219 | 0.199 | 0.002 |

**Figure 2:**
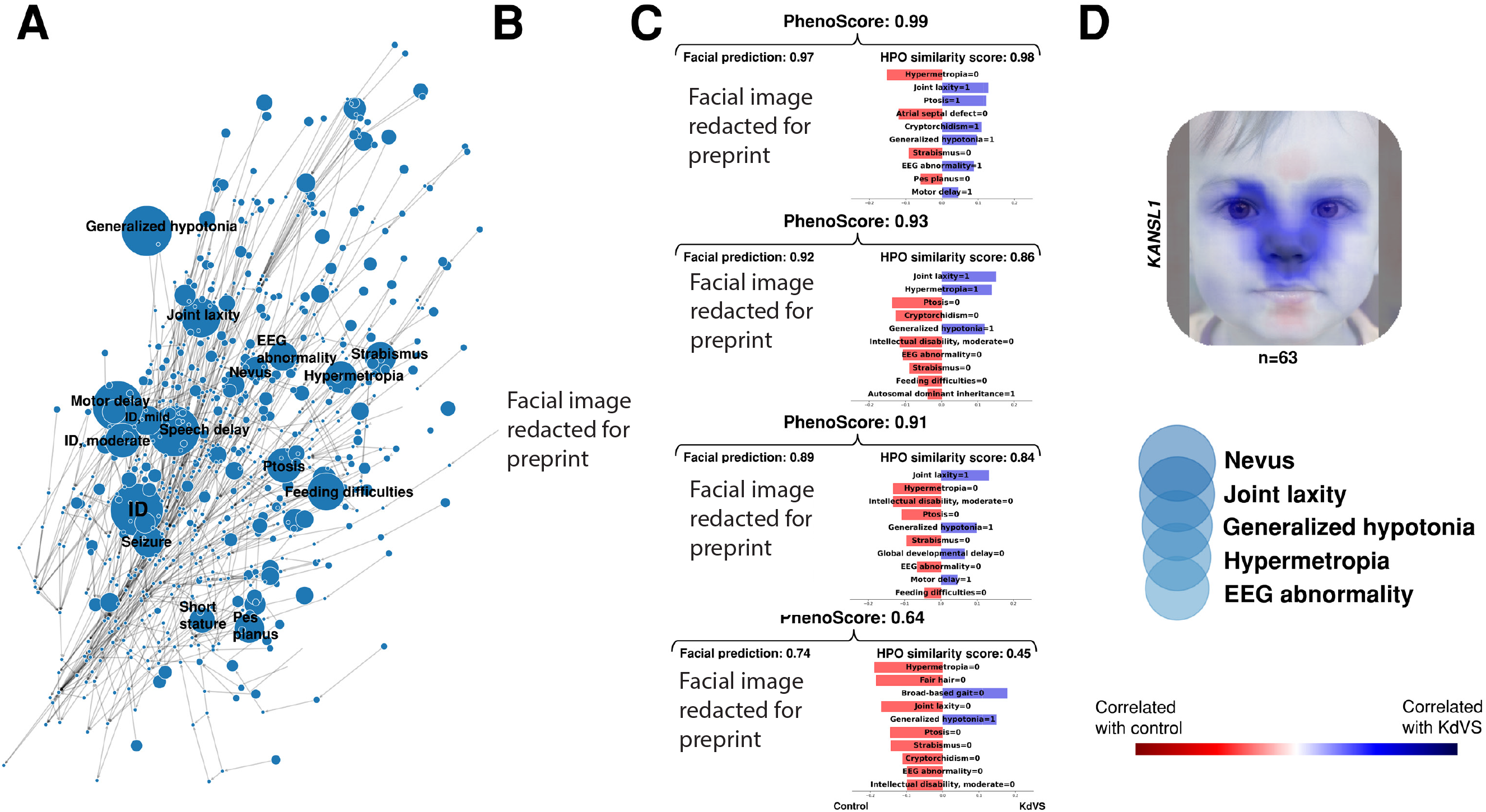
A) The HPO terms of all included individuals with Koolen-de Vries (KdVS) are shown here. HPO terms present in 20% or more of the individuals are annotated with text, and larger nodes correspond to a higher prevalence of that specific clinical feature. ID = intellectual disability. B) Four individuals diagnosed with Koolen-de Vries syndrome are presented here (written informed consent for the publication of these facial images was obtained). These were randomly selected from the included dataset without any selection criterion. C) For the four randomly selected individuals, three predictions are shown: using the facial image, using the phenotypic data, and finally, the PhenoScore, which combines both. Furthermore, heatmaps are generated using local interpretable model-agnostic explanations (LIME) to see which facial areas are most important according to our model, where blue correlates with KdVS and red areas correlate with controls. The nose and eyes are clearly prioritized, corresponding to the known dysmorphic features in Koolen-de Vries. Furthermore, the most important clinical features are shown for each individual and the contribution (corresponding to the LIME regression coefficient) of that feature to the prediction. D) Finally, a summarized heatmap was generated to investigate the overall most important facial and phenotypic features. We averaged the heatmaps of the five individuals with Koolen-de Vries with the highest prediction. Next to that, to obtain the most important clinical features, too, we averaged the LIME regression coefficient for the different symptoms of the five highest-scoring individuals based on HPO. Shown clinical features are ordered based on importance, and the size of the circle indicates the relative importance of the feature.

We next randomly excluded four individuals (facial images shown in Figure 2) from the training dataset and retrained PhenoSscore, allowing us to test the performance of PhenoScore when treating them as if diagnoses of KdVS were unknown. We then used PhenoScore to predict the similarity of these four individuals when comparing them with 59 remaining individuals with KdVS in the training set. PhenoScore output was displayed using LIME, providing heatmaps of prioritized facial information according to PhenoScore (Figure 2). In addition, the most important clinical features according to PhenoScore to be predictive for KdVS were summarized by numerically scoring and ranking them. According to PhenoScore, the nose and eyes are the most important facial parts when recognizing KdVS — while the presence of nevi, joint laxity, hypotonia, hypermetropia, and EEG abnormalities are the clinical features of interest. This is completely consistent with expert opinion and the literature [51–53] and shows that the prediction is based on the extracted facial features from 2D photos and phenotypic data in HPO — harnessing the power of both and outperforms the separate predictions.

### 2.3 Expanding PhenoScore to 26 syndromes

After our proof-of-concept using KdVS, we next assessed the performance of PhenoScore for the classification of other genetic syndromes too. Hereto, we selected 25 syndromes (Table 1 and Supplemental Table 1) including both clinically well-recognizable syndromes based on facial gestalt, such as Kleefstra syndrome (OMIM #610253, caused by pathogenic variants in *EHMT1*), Helsmoortel-van der Aa syndrome (OMIM #615873, caused by pathogenic variants in *ADNP*) and Coffin-Siris syndrome (OMIM #135900, *ARID1B*), but also more recently identified syndromes for which facial gestalt is less prominent, including IDDAM (OMIM #615032, *CHD8*) and IDDFBA (OMIM #618089, *FBXO11*).

Analyzing all these syndromes, we demonstrate that PhenoScore is a statistically significant improvement on using either feature set alone, and therefore, the whole is more than the sum of its parts in this case (median Brier score 0.22 for facial features on the whole dataset, 0.16 for HPO data and 0.15 for PhenoScore, *p <*0.001; median AUC 0.71 for facial features, 0.85 for HPO data and 0.87 for PhenoScore, *p <*0.001, Table 1). Furthermore, our post hoc checks show that there was no overfitting using the internal control dataset (see Supplemental Table 2).

For 25 of 26 syndromes (96%), PhenoScore was able to identify predictive features that characterized these syndromes and recognized a distinct phenotypic entity (Table 1). As expected, and visualized in the LIME heatmaps (Figure 3), these features corresponded remarkably well with those described in the literature. For instance, for Helsmoortel-van der Aa syndrome (OMIM #615873), the facial- and forehead regions are prioritized in the predictions, as seen in the generated heatmap (Figure 3d) — corresponding with the known dysmorphic characteristics for this syndrome.

**Figure 3:**
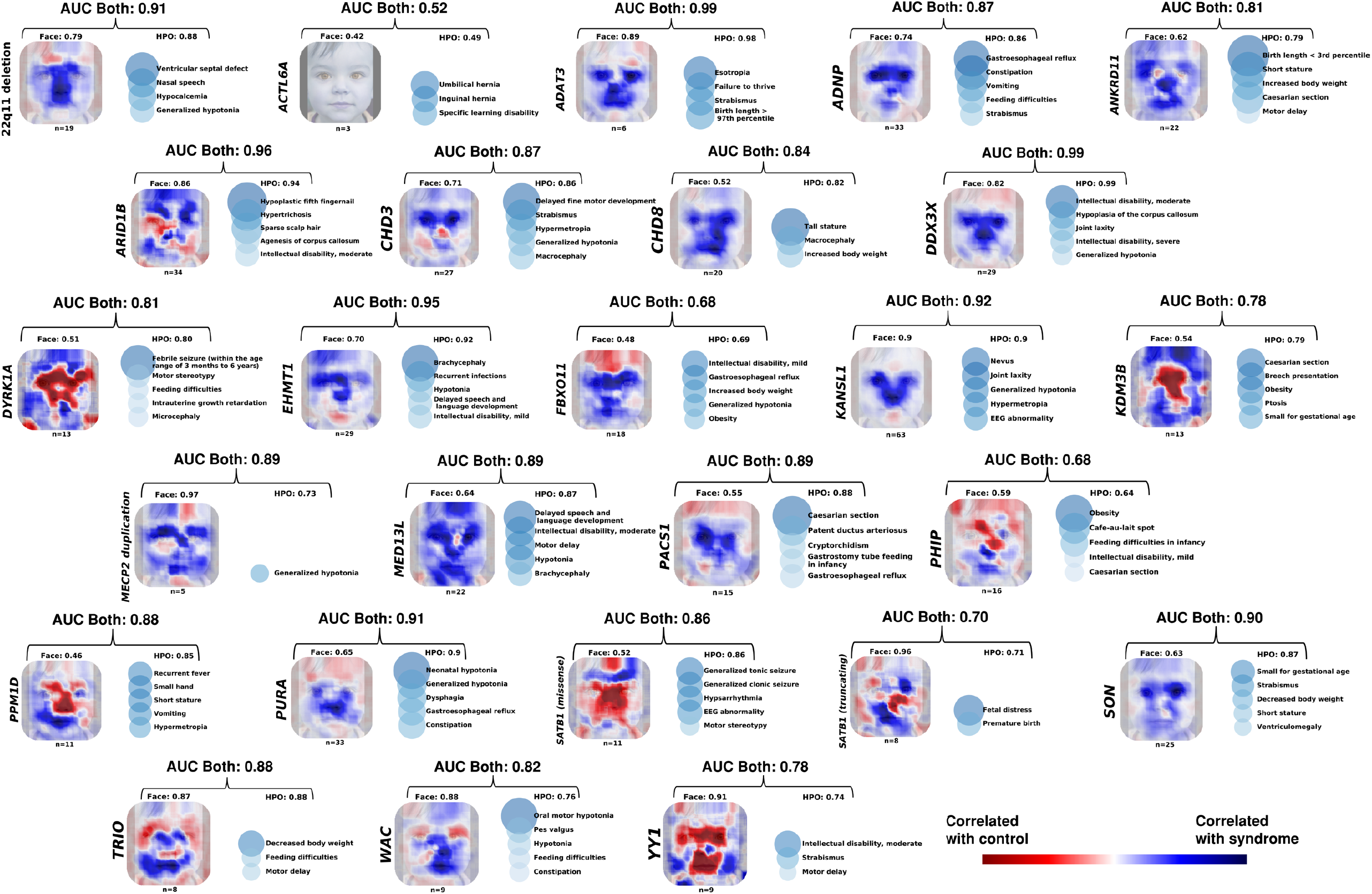
The heatmaps and most important clinical features of all 26 genetic syndromes included in this study are displayed in this figure. The facial heatmaps are the average LIME heatmaps of the five individuals per genetic syndrome with the highest predictive score based on the facial data alone. For the phenotypic data, the positive LIME regression coefficients per symptom were averaged of the top-scoring five individuals based on the phenotypic data. The standard face used as background is a non-existent person generated using StyleGAN. [74].

Moreover, for a genetic syndrome which lacks explicit facial features, like IDDAM, apparent overgrowth symptoms, such as macrocephaly and tall stature, were identified as significant predictors, while no relevant facial features were extracted, as displayed in the heatmap and summarized ranking scores. A similar case is made for the genetic disorder associated with pathogenic variants in *DYRK1A*: while the classifier based only on the facial features does not provide any meaningful predictions, the addition of other phenotypic data in HPO did allow PhenoScore to distinguish this syndrome as a phenotypic entity. These data suggest that PhenoScore objectively extracts, distinguishes, and visualizes the specific clinical features for genetic syndromes and highlights that the addition of non-facial phenotypic data in HPO is essential.

### 2.4 PhenoScore is scalable as it requires only a low number of individuals for training

Most genetic disorders are individually rare, with sometimes only 3-5 individuals reported world-wide. We therefore next investigated how many data sets PhenoScore requires for accurate classification of a specific syndrome. We checked the performance of PhenoScore while increasing the number of individuals in the complete dataset of 26 genetic syndromes with the combination of facial- and HPO features, starting with only 2 individuals. This analysis revealed that, with three individuals to train on, the median classification performance for the investigated syndromes is already clinically acceptable (AUC 0.85; Figure 4). The classification performance can be further improved when the training sets increase in size (median AUC 0.90 with seven individuals, 0.95 for 17 individuals).

**Figure 4:**
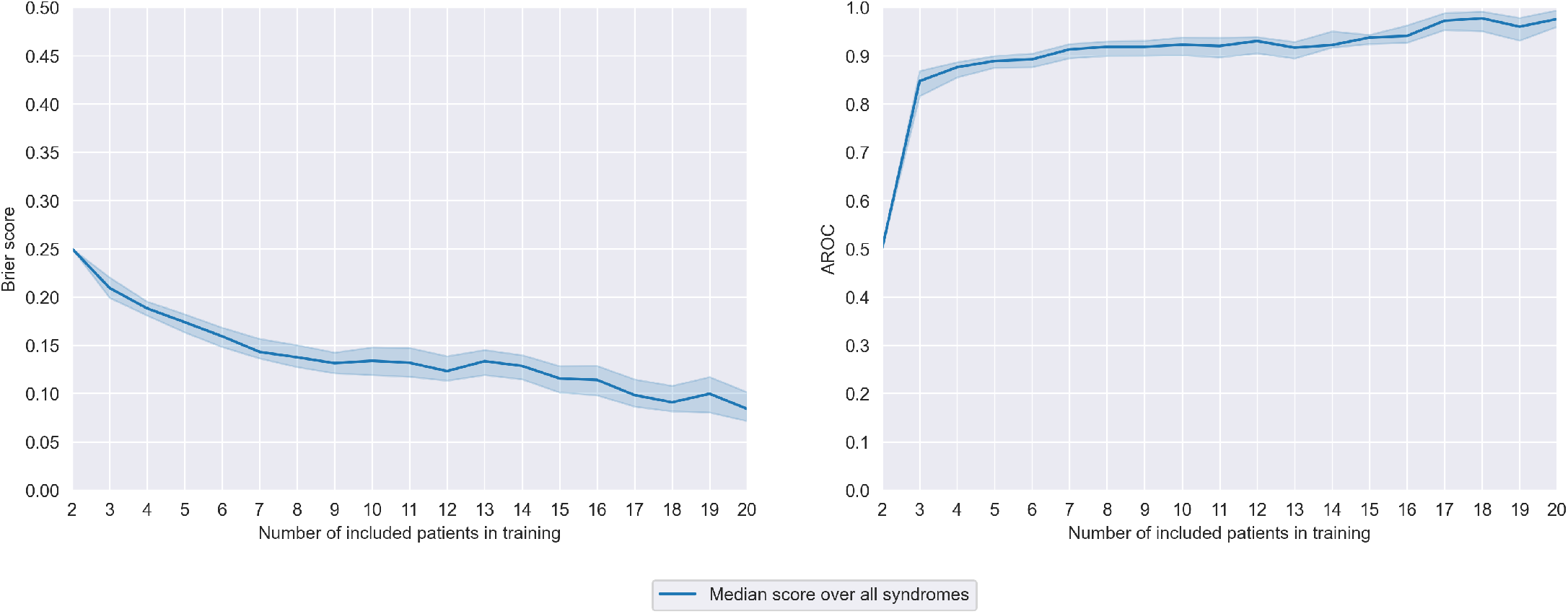
The performance of the SVM using both facial- and HPO features with different sizes of the training set is shown here. Both the median Brier score and the median AUC improve if the number of individuals to train on is larger — as would be expected. Interestingly, only three individuals are needed for an already acceptable classification performance.

### 2.5 Use case 1: Objective clinical quantification for the interpretation of molecular VUS

To display the power of PhenoScore in the clinical interpretation of variants at an individual level, we reassessed reported VUSs (ACMG class 3) in the Radboudumc department of Human Genetics. These individuals were not included in the training of PhenoScore and can therefore be considers real out-of-sample cases. In total, we identified 15 individuals in whom a class 3 variant was reported in either of 11 of the 26 syndromes (Supplemental Table 3). PhenoScores were calculated, and when using thresholds of ≤0.30 (for ‘no phenotypic match’) and ≥0.70 (for ‘phenotypic match’), PhenoScore was able to classify 9/15 (60%) of the cases as either match (*n*=2) or no match (*n*=7). The other 6 cases had an inconclusive PhenoScore result (scores *>*0.30 but *<*0.70). Interestingly, for only 1/9 cases for which PhenoScore was conclusive, the clinician made a decision for the VUS based on the phenotype — PhenoScore was essential for the other eight cases. Importantly, parallel functional follow-up for 4 variants confirmed the PhenoScore outcome, whereas for the remaining cases, functional follow-up was inconclusive.

### 2.6 Use case 2: Next-generation phenomics for the generation of sophisticated genotype-phenotype correlations

Genotype-phenotype studies for rare diseases are often performed to gain insight into the clinical spectrum, which allows clinicians to provide a more accurate counseling of individuals with rare diseases. Molecularly, the toolkit to gain in-depth insight into aspects of pathogenicity is generally applied in a research setting, and thus often not readily available for diagnostic follow-up. From a clinical perspective, analyses are often limited to cluster analysis and/or principle component analysis, but without being able to determine what aspects clinically distinguish subtypes, if identified. We tested whether PhenoScore can improve these hypothesis-driven approaches to distinguish, or discover, clinical subtypes.

For two genetic syndromes in our dataset, i.e. *SATB1*-associated neurodevelopmental disorders (OMIM #619228)[54], and Helsmoortel-Van Der Aa Syndrome (OMIM #615873, caused by disruption of *ADNP* [55]), it has previously been determined that there are (at least) two molecular-subtypes. For *SATB1*, it has also been acknowledged that individuals with missense variants and those with loss-of-function variants, are clinically different. As proof-of-concept, PhenoScore convincingly distinguished two groups for *SATB1* (Brier score 0.18, AUC 0.81, *p* = 0.02), confirming the original results [54]. For *ADNP*, it was recently shown that individuals with pathogenic variants in *ADNP* show one of two distinct methylation signatures (type 2, when variant affects position between c.2000 and c.2340; or type 1, when the variant occurs outside of this interval), suggesting the possibility of two syndromes associated with this gene [56]. Clinically, however, these individuals could not be conclusively distinguished [57]. Prior to determining PhenoScores, we categorized the individuals as having either a type 1 or type 2 *ADNP* signature. Initially, we assessed the performance of PhenoScore using only individuals (*n*=33) for whom both facial photographs and clinical features were available, but failed to identify a statistically significant difference between the groups (Brier 0.30, AUC 0.52, *p* = 0.35). However, using the *ADNP* Human Disease Gene website, we could collect HPO-only data of more individuals. Using this dataset, we obtained clinical features in HPO of 58 individuals (29 in each group), and on these data PhenoScore did show evidence for two phenotypically different entities (Brier 0.24, AUC of 0.71, *p* = 0.02). Inspecting the generated PhenoScore explanations for clinically relevant differences (Figure 5), it seems that recurrent infections and gastrointestinal problems (reflux, constipation, feeding difficulties) are 2-3 times more common in type 2 than in type 1.

**Figure 5:**
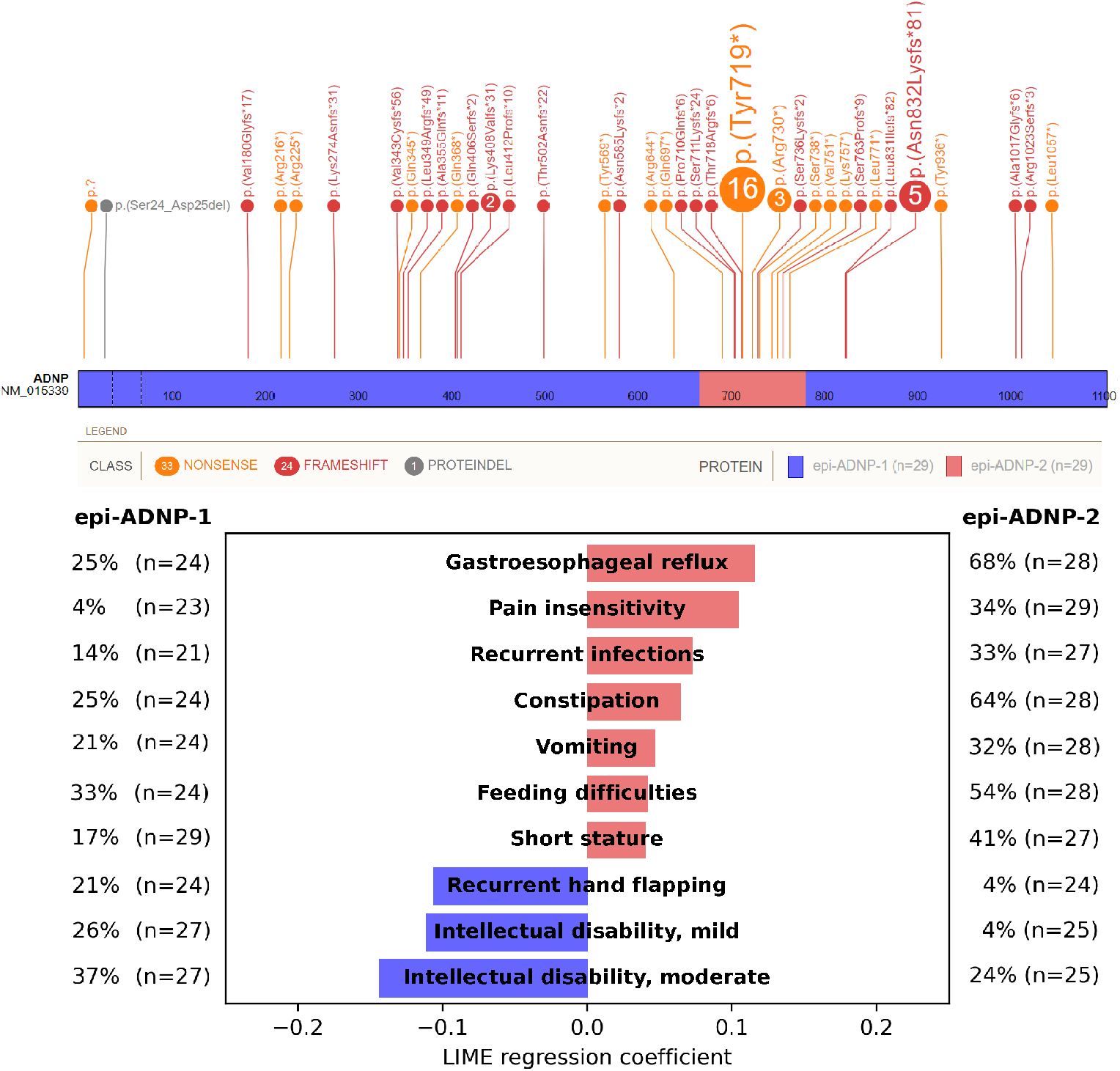
Above: a lollipop plot (generated using St. Jude’s ProteinPaint) of the genetic variants currently collected using the *ADNP* HDG website [62]. Of the 58 included individuals, 29 had a variant in the c.2000-2340 region, indicated by others as having a different methylation signature than variants outside this [56]. Using only the HPO module of our PhenoScore framework, we first matched the groups on gender-, ethnicity- and age when possible to create two groups of the same size (29 vs. 29). We then trained a classifier on the two groups and found a significant difference (Brier score of 0.24, AUC of 0.71, *p* = 0.01). Below: the most important clinical features according to our model (determined using LIME) and the corresponding prevalence in both groups.

## 3 Discussion

PhenoScore provides a significant step in the advancement of AI in clinical genetics: a novel machine learning phenomics framework unifying facial and phenotypic features using high-quality data directly from affected individuals instead of generic phenotypic descriptions of a syndrome. Others have introduced AI in this domain of healthcare, with for instance the application of using HPO terms to prioritize genetic variants while comparing individuals to the known phenotype of disorders in the literature [30, 31, 36, 58]. The utilization of facial recognition technology to assist clinicians in diagnosing individuals has been successful too, with most, unfortunately, relying on proprietary commercial algorithms [35, 37–42]. We now show a next step, with an open-source framework that takes the complete phenotype into account, including both facial- and phenotypic features directly from affected individuals, and uses AI to provide a score on how well the patient’s phenotype (as a whole) matches individuals with a known syndrome.

PhenoScore detected a recognizable phenotype in all but one investigated genetic syndrome (25/26; 96%), and only needed as little as three individuals for classification performance. In this manner, PhenoScore assists clinicians and molecular biologists in quantifying phenotypic similarity, at both an individual- and group level for theoretically all OMIM-listed disorders. The sole disorder for which PhenoScore failed to identify a phenotype was for variants in *ACTL6A*. Interestingly, this is the only of 26 syndromes that has not been recognized by OMIM as a genetic disorder, due to lack of (phenotypic) evidence in combination with the fact that individuals in whom the *ACTL6A* variants were uncovered, had not been tested by trio whole exome sequencing. OMIM therefore concluded that it is uncertain that the variants in *ACTL6A* cause the phenotype of these individuals.

Assisting variant classification of VUSs is an obvious use-case for PhenoScore. Of course, several *in vitro* functional assays are available to assess variant pathogenicity, but so far these are mostly used for genes involved in oncogenetic disorders [59, 60]. For neurodevelopmental disorders, these assays are scarce since they need to be developed on a gene-per-gene basis, and for these rare disorders, this is usually not cost-effective and solely done for research purposes. Other methods to assess genetic variants include protein structural analysis [61], which however still relies on the availability of relevant protein structures. Our approach theoretically works for any (genetic) condition with a recognizable phenotype, provided there are sufficient individuals for training the algorithm, and that HPO data and 2D-facial photos are available. Indeed, PhenoScore is as good as its input data. In the field of rare diseases, however, major efforts are put in obtaining these high-quality quantitative phenotypic data, as for instance shown by collections of datasets by the Human Disease Gene website series [62], GeneReviews, DECIPHER and OMIM [63–65]. Here, the use of HPO terms, in combination with the use of Resnik scores (ensuring that the use of similar HPO terms leads to comparable results), minimizes the effect of variation in clinical terminology used between clinicians, and thus deriving the most benefits from the AI-based quantitative phenotypic framework.

PhenoScore also helped to objectively obtain genotype-phenotype correlations, by training on suspected phenotypic subgroups combined by permutation testing to quantify statistical significance. We replicated earlier findings in *SATB1*, quantitatively underscoring that truncating variants lead to a significantly different phenotype than missense variants [54].Whereas for SATB1 the different phenotypes were also subjectively identifiably from expert opinion, the power of PhenoScore was shown by demonstrating the existence of two distinct phenotypes associated with Helsmoortel-van der Aa syndrome. Molecularly, two different methylation signatures have been published, which were discriminated by the mutation location in *ADNP* [55–57]], but for which clinically, no differences were observed. PhenoScore was not only able to prove the existence of clinically distinctive groups, but also provided insight into which clinical features separates the two clinical entities. For instance, neurodevelopmental problems are more common in the *ADNP*-type 1, while gastrointestinal symptoms, recurrent infections and short stature are 2-3 times more common in *ADNP*-type 2. These clinical features have a significant impact on an individual’s quality of life, hence, by identifying these subgroups, PhenoScore directly impacted clinical recommendations for these individuals and families.

These subgroup analyses could in theory be performed for every (genetic) syndrome caused by different types of SNVs or CNVs — which is the case in a significant portion of the currently ∼1600 known NDD genes. While recognizing specific novel subgroups is a first step towards personalized medicine and provides improved clinical prognosis and recommendations (as shown for the subgroups in *ADNP* and *SATB1*), not finding a distinct difference is useful too: it helps to assess whether two types of genetic variation have the same effect (i.e. whether missense variants actually cause haploinsufficiency). Furthermore, PhenoScore could be utilized to find phenotypic outliers, of whom the molecular mechanism leading to disease might be novel. By quantifying the complete phenotypic similarity and visualizing differences between (sub)groups, PhenoScore empowers detailed genotype-phenotype studies, leading to new insights on both the genetic- and phenotypic level.

The discriminating clinical features for the two *ADNP*-related disorders were not represented in a different facial gestalt, emphasizing the importance of adding HPO data across all organ systems. In addition, given that these two phenotypic subgroups were not identified from more subjective clinical analysis, using a predefined structured AI method of phenotypic data analysis provides novel insights. To facilitate easy use in routine clinical care, it is, however, also of paramount importance to be able to intuitively understand the AI output. We therefore also provided graphical output such as facial heatmaps to visualize which (facial) features specified PhenoScore output.

Detailed genotype-phenotype analysis could in theory be performed for every (genetic) syndrome, suggesting that PhenoScore may be a valueable tool to also foster novel molecular insights. That is, for many of the 1,600 known genes associated to an NDD phenotype, multiple types of genetic variants (e.g. SNVs and CNVs) may cause the disorder. Whereas the molecular mechanism for CNVs often relate to dosage-sensitivity, such as haploinsufficiency, the mechanisms for SNVs leading to missense variants in those genes, are often less pertinent. PhenoScore may assess phenotypic differences between individuals with the same syndrome, but caused by either CNVs (‘group 1’) or missense variants (‘group 2’) and help to establish whether those missense variants are also haploinsufficient. Similarly, PhenoScore could be utilized to find phenotypic outliers, of which the molecular mechanism leading to disease might be novel.

In conclusion, PhenoScore bridges a gap between the fields of AI and Clinical Genetics by quantifying phenotypic similarity, assisting not only in genetic variant interpretation, but also facilitating objective genotype-phenotype studies. We showcased its use for individuals with NDD, whose phenotypes were captured using HPO. PhenoScore can, however, also easily be used beyond the field of rare disease, as adjustments to use other (graph-based) ontologies, such as for instance SNOMED [66], can readily be integrated. The PhenoScore AI-based framework is thus easily extended to other domains of (clinical) genetics, or even to completely different branches of medicine, due to its open-source modular design.

## 4 Materials and Methods

### 4.1 Inclusion of individuals

The literature was searched for clinical studies which included facial photographs for 26 randomly selected genetic syndromes associated with NDD. The photographs were collected and clinical features, if available, were converted to HPO terms. Currently, PhenoScore is trained using data of 501 non-familial individuals diagnosed with one of the 26 different genetic syndromes, collected from 81 different publications (see Table 1 for the complete overview of the demographics per genetic syndrome and Supplemental Table 1 for all publications used as sources for the data used in this study). The phenotypic data were uploaded to the specific gene website in the HDG website series [62] to ensure their public availability. The use of these data was approved by the ethical committee of the Radboud university medical center (#2020-6151).

### 4.2 Data processing

To obtain a representative control group for our machine learning models, for each syndrome with *n* individuals, *n* age-, sex- and ethnicity matched controls with a neurodevelopmental disorder seen at our outpatient clinic at the Radboud university medical center were selected as described previously [39] from our internal control database with over 1200 individuals with both facial image and quantitative phenotypic data available (for a complete overview of the workflow of this study, please see Figure 1). When no matched control was available, that particular individual was excluded from our analysis. Next to that, when individuals were related to each other, one individual was chosen (based on the quality of the picture) from that family. For each syndrome, cross-validation was used to assess the performance of the classifiers. The number of folds during the cross-validation procedure varied due to the considerable variation in dataset size: for every syndrome with at least ten individuals, 5-fold cross-validation was used — otherwise, leave-one-out cross-validation was chosen. As the selection of the randomly selected controls might significantly influence the performance, for each genetic syndrome, different controls were sampled during ten random restarts and the mean AUC and Brier scores of these ten iterations were noted. Furthermore, to confirm the source of the data did not significantly influence our results, we did post hoc checks by using not only the individuals from our internal control dataset, but the other included syndromes as well as controls.

### 4.3 Extraction of facial features

The facial features were extracted using VGGFace2 [67, 68], a state-of-the-art facial recognition method that utilizes a deep neural network. To avoid overfitting, we did not retrain VGGFace2, but used its pretrained weights instead. The images were then processed by VGGFace2 and the representation in the penultimate layer of the network was obtained. This representation was then used as the facial feature vector. The process was performed as described previously: for the entire (technological) methodology, please see [69].

### 4.4 Phenotypic similarity

To create a homogeneous dataset, the phenotype of every individual in this study was manually converted into HPO terms [14]. A selection of HPO terms and all their child nodes were removed to eliminate any subjectivity in assessing an individual. These were *Behavioral abnormality (HP:0000708), Abnormality of the face (HP:0000271), Abnormal digit morphology (HP:0011297), Abnormal ear morphology (HP:0031703), Abnormal eye morphology (HP:0012372)*, and every node which is a child node of either of these. We chose these terms as these are either facial features (to be assessed by our facial recognition model) or are suspected to vary across clinicians doing the assessment of an individual. In this manner, 3 810 HPO terms were excluded with 12 259 terms remaining. To further reduce possible inter-observer variability, the phenotypic similarity between individuals was calculated using the Resnik score [70], since it takes the semantic similarity between symptoms into account. The Resnik score utilizes the information content (IC) of a symptom. In an ontology akin to the HPO, the IC of a specific term can be seen as a measure of the rarity of a term. Naturally, terms closer to the root of the HPO tree have a lower IC. For instance, *Abnormality of the nervous system (HP:0000707)* has an IC of 0.60. In contrast, *Focal impaired awareness motor seizure with dystonia (HP:0032717)*, significantly further down the HPO tree, has an IC of 8.97. This corresponds to our intuition: rare features provide more information than common features — since the prior probability of an individual reporting a rare symptom is, by definition, smaller. The Resnik score uses this property by defining the similarity between two HPO terms as the IC of their most informative (that is, with the highest IC) common ancestor in the HPO tree. Since terms lower in the tree have a higher IC, the most informative common ancestor corresponds to the last HPO term, which has both compared HPO terms as child nodes when traversing the tree downwards. As an example: for the HPO terms *Reflex seizure (HP:0020207)* and *Focal motor seizure (HP:0011153)*, the most informative common ancestor is *Seizure (HP:0001250)*, which has an IC of 1.70. The Resnik similarity score for *Reflex seizure (HP:0020207)* and *Focal motor seizure (HP:0011153)* is therefore 1.70. Next, we used the best-match average (BMA) to calculate the similarity between two individuals (who usually report multiple HPO terms), in which the average is taken over all best-matched pairwise semantic similarities, as previous studies determined it to be most effective [71]. The idea is similar to that discussed above: if two individuals share a rare symptom *(Focal impaired awareness motor seizure with dystonia (HP:0032717)*, for instance), they are more similar than two individuals who only share a common symptom such as *Abnormality of the nervous system (HP:0000707)*. The Resnik similarity score was calculated for every individual and control and then averaged for both groups. In the end, this led to a *n*x2 matrix for the HPO features: an average similarity score for each individual versus affected individuals and a score for each individual versus the control group. We calculated the BMA Resnik score between the individuals using the phenopy library in Python 3.8 [72].

### 4.5 Construction of machine learning model

Finally, the data were used to train a binary classifier. We selected a support vector machine (SVM) as our classifier, known for its excellent overall performance in classification tasks. The SVM was trained using the standard radial basis function kernel and a hyperparameter grid search for C.

After determining the predictive performance of the model, we determined how many data the classifier needed for an acceptable classification performance in clinical practice. Per syndrome, we started with randomly selecting two individuals and two matched controls, training the model on those, and using the rest of the individuals (*n −* 2, as one individual is used as training data) and matched controls as a test set (two individuals that were not used in the first iteration as the grid search in the SVM classifier needs at least two training samples). We ran ten random restarts, randomly selecting another individual and matched control in each iteration. In each restart, leave-one-out cross-validation was employed. The Brier score and AUC were noted and averaged over the ten restarts. Next, the size of the training set was increased by one patient, and one matched control, still using the rest of the individuals (now *n −* 3) and matched controls as the test set. By increasing the training set by one individual and matched control each time and recording the performance, the model’s performance with an increasing number of individuals is assessed.

The Wilcoxon signed-rank test was used to determine statistically significant differences in the performance of the classifiers since it is a non-parametric test and, therefore, suitable — as these data are not normally distributed.

### 4.6 Explainability of predictions

To see which features contained important information for our model, we generated Local Interpretable Model-agnostic Explanations (LIME) [48, 49]. The main idea of this method is to train a relatively simple local surrogate model to approximate the predictions of the model of interest. Next, the original input data is perturbed, and the corresponding change in predictions is inspected to obtain the relative importance of individual features. A key advantage of LIME is that it is applicable to any model and can therefore be used directly on top of our pipeline.

When using LIME for image data, it is common practice to divide the image into several segments, called superpixels. Therefore, we generated a raster of 25*×*25 pixel squar**e**s for each facial image, randomly offset for each of 100 runs. Each pixel’s relative importance was averaged over these runs to obtain a higher resolution visualization of their significance. For the clinical data, the original HPO features were perturbed to obtain the most significant ones in predictions. In this case, LIME uses input data in which some HPO features are added and some are removed from the input data, to see what the effect on the prediction is.

LIME explanations were generated for the individuals with the five highest predictions scores. These explanations were then averaged, to obtain an overall explanation representative for that specific genetic syndrome. To ensure only real important features were recovered, only HPO terms that were identified in at least three individuals were used in this analysis.

### 4.7 Hypothesis testing

To see whether we could extend the use of our classifier to other applications than the reclassification of VUSs, we designed a random permutation test for the performance of our model. This enables the testing of a specific hypothesis for facial features, phenotypes, or both. An example would be determining whether a newly discovered genetic syndrome consists of several (phenotypic/facial) subtypes. Using our framework, we trained a classifier on the labels of the suspected subgroups. By performing a random permutation test, a *p*-value is calculated, so that the appearance of the subgroups can be quantified. For a complete overview of the exact methodology of this permutation test, please see the Supplemental Methods.

## Data Availability

All data produced in the present study are available upon reasonable request to the authors

https://github.com/ldingemans/PhenoScore

## Data and code availability

The code of PhenoScore created during this study is freely available at https://github.com/ldingemans/PhenoScore, to enable anyone to apply PhenoScore to their own dataset. Included in PhenoScore are two examples: the data for the *SATB1* subgroups (positive example) and random data (negative example). The used dataset in this study is not publicly available due to both IRB and General Data Protection Regulation (EU GDPR) restrictions since the data might be (partially) traceable. However, access to the data may be requested from the data availability committee by contacting the corresponding author.

## Acknowledgements

We are grateful to the Dutch Organisation for Health Research and Development: ZON-MW grants 912-12-109 (to B.B.A.d.V. and L.E.L.M.V.), Donders Junior researcher grant 2019 (B.B.A.d.V. and L.E.L.M.V.) and Aspasia grant 015.014.066 (to L.E.L.M.V.). The aims of this study contribute to the Solve-RD project (to L.E.L.M.V.), which has received funding from the European Union’s Horizon 2020 research and innovation program under grant agreement No 779257. R.F.K acknowledges financial support of the Research Fund of the University of Antwerp (Methusalem-OEC grant – “GENOMED”). The work of G.J.L. is supported by New York State Office for People with Developmental Disabilities (OPWDD) and NIH NIGMS R35-GM-133408.

## Author information

Conceptualization: A.J.M.D, M.H, L.E.L.M.V, B.B.A.d.V, M.A.J.v.G; Data curation: A.J.M.D, K.M.G.T, L.G, J.v.R, N.d.L, J.S.H, R.P, I.J.D, E.d.B, J.d.H, J.v.d.S, S.J, B.W.v.B, N.J, A.T.V.v.S, T.K, D.A.K, F.K, H.V.E, G.J.L, F.S.A, A.R, R.M, D.B, P.J.v.d.S., G.S, L.E.L.M.V, B.B.A.d.V; Formal Analysis: A.J.M.D, M.H; Funding acquisition: L.E.L.M.V, B.B.A.d.V; Investigation: A.J.M.D, M.H; Modelling: A.J.M.D, M.H.; Software development: A.J.M.D; Writing – original draft: A.J.M.D, M.H, L.E.L.M.V, B.B.A.d.V, M.A.J.v.G; Writing – review and editing: all authors.

## Ethics declaration

In this study, data from the Biobank ‘Intellectual Disability’, which is part of the Radboud Biobank initiative (for more information, see [73] or https://www.radboudumc.nl/en/research/radboud-technology-centers/radboud-biobank) were used. Within this biobank, phenotypic and molecular data have been systematically captured for individuals with (non-)syndromic ID referred to the Radboud university medical center. The use of this dataset was approved by the ethical committee of the Radboud university medical center (#2020-6151). Furthermore, the authors declare no competing interests.

## 5 Supplementary data

### 5.1 Supplementary methods

#### 5.1.1 Permutation test for hypothesis testing

To provide a *p*-value for our classification results and to enable the use of our framework for the recognition of specific (sub)groups in genetic syndromes, we developed a permutation test — inspired by the test described by Lopez-Paz & Oquab [75]. For every group of individuals of interest, a group of age-, sex- and ethnicity-matched controls is sampled from our control database. We extract the facial features using VGGFace2 and calculate the HPO similarity using the Resnik score, after which a SVM is trained using cross-validation. A grid search for the optimal hyperparameters is performed, and the Brier score is calculated for this combination of the group of individuals of interest and the matched controls. This process is precisely the same as in our standard analysis. Next, we randomly permute the labels (here, the labels correspond to whether an individual has the syndrome or is a control) 100 times. We ensure that the number of positive and negative classes is the same as in our original distribution of the labels. For each permutation, we repeat the process of training a SVM to obtain a Brier score. We then perform a one-sided Mann-Whitney U test to quantify the probability of the classification results being statistically significantly smaller (since it is the Brier score we are comparing) than the randomly permuted scores.

To further strengthen our permutation test, we repeat the process five times in total, randomly sampling matched controls from our database in each repetition. The five obtained *p* values were then combined using Fisher’s method [76] to gather a definitive *p*-value for this classification task and, therefore, for this specific group of individuals of interest.

For the analyses for both *SATB1* and *ADNP*, we do not need to sample controls from our database. However, these datasets are usually imbalanced, sometimes leading to problems for the classifier. We therefore undersample the majority class to the size of the minority class by matching the individuals on ethnicity, sex and age (in that order) and increase the number of permutations of the labels to 1 000 (since we cannot repeatedly sample controls).

Finally, to have a negative control group for this test, we randomly sampled individuals from our control database and calculated *p*-values for those. We did this for different cohort sizes (*n* = 3, 5, 10, 20 and 40) for in total 50 trials. Of those 50 trials, two resulted in a *p*-value smaller than 0.05 - exactly what would be expected by random chance in this number of trials. This shows that our approach leads to the to-be-expected number of false alarms.

**Supplementary table 1:** A list is shown of the used publications per syndrome to create the dataset by extracting the phenotypic data and photographs of individuals in these papers. For several syndromes, not (yet) published individuals were added to the dataset, as indicated by Not published.

| Genetic syndrome | PMID of used publications |
| --- | --- |
| 22q11 deletion | 15831592,17041934,18636631,1956057,21200182,25317860,3816857,12548732 [77-84] |
| ACTL6A | 28649782 [85] |
| ADAT3 | 23620220,26842963 [86, 87] |
| ADNP | 24531329,25217958,27031564,29724491 [55, 88-90] |
| ANKRD11 | 19920853,21527850,21654729,22307766,23494856,26269249 [91-96] |
| ARID1B | 19034313,22405089,22426309,22585544,23906836,24569609,26395437,26754677,27112773,28323383,30055038,30349098,31981384,32339967 [97-110] |
| CHD3 | 30397230,32483341 [111, 112] |
| CHD8 | 23160955,24998929,31001818,31721432,Not published [113-116] |
| DDX3X | 26235985 [117] |
| DYRK1A | 18405873,21294719,23099646,23160955,25707398,Not published [113, 118-121] |
| EHMT1 | 22670141,Not published [122] |
| FBXO11 | 27479843,28343630 [123, 124] |
| KANSL1 | 16906164,17601928,18628315,21094706,22544363,26306646,Not published [51, 53, 125-128] |
| KDM3B | 30929739 [129] |
| MECP2 duplication | 18854860,18985075 [130, 131] |
| MED13L | 23403903,24781760,25712080,28645799,29511999,29959045 [132-137] |
| PACS1 | 26842493 [138] |
| PHIP | 29209020 [139] |
| PPM1D | 28343630 [124] |
| PURA | 27148565,29097605,29150892 [140-142] |
| SATB1 (missense + truncating) | 33513338 [54] |
| SON | 24896178,27256762,27545680 [4, 143-145] |
| TRIO | 26721934,27418539 [146, 147] |
| WAC | 23033978,26264232,26757981 [2, 148, 149] |
| YY1 | 21076407,28575647 [1, 150] |

**Supplementary table 2:** The Brier scores of the support vector machine (SVM) classifier are displayed here, now with the other individuals included in this study as the control dataset, instead of the controls from the Radboud university medical center. The results are slightly worse than on the RUMC control dataset, as expected, since not for not every individual, a control is available because the RUMC control dataset is significantly larger than the number of individuals included in this study.

| Genetic syndrome | Facial data only | HPO data only | PhenoScore |
| --- | --- | --- | --- |
| 22Q11 | 0.224 | 0.120 | 0.120 |
| ACTL6A | 0.248 | 0.231 | 0.219 |
| ADAT3 | 0.165 | 0.073 | 0.067 |
| ADNP | 0.226 | 0.187 | 0.178 |
| ANKRD11 | 0.253 | 0.148 | 0.146 |
| ARID1B | 0.175 | 0.100 | 0.091 |
| CHD3 | 0.222 | 0.154 | 0.154 |
| CHD8 | 0.239 | 0.160 | 0.126 |
| DDX3X | 0.226 | 0.022 | 0.024 |
| DYRK1A | 0.256 | 0.182 | 0.159 |
| EHMT1 | 0.191 | 0.124 | 0.107 |
| FBXO11 | 0.273 | 0.218 | 0.224 |
| KANSL1 | 0.122 | 0.122 | 0.105 |
| KDM3B | 0.283 | 0.167 | 0.161 |
| MECP2 | 0.167 | 0.252 | 0.199 |
| MED13L | 0.250 | 0.186 | 0.177 |
| PACS1 | 0.277 | 0.197 | 0.198 |
| PHIP | 0.198 | 0.141 | 0.139 |
| PPM1D | 0.255 | 0.125 | 0.116 |
| PURA | 0.202 | 0.118 | 0.126 |
| SATB1 (missense) | 0.263 | 0.119 | 0.113 |
| SATB1 (truncating) | 0.137 | 0.205 | 0.186 |
| SON | 0.237 | 0.135 | 0.121 |
| TRIO | 0.132 | 0.178 | 0.173 |
| WAC | 0.137 | 0.193 | 0.171 |
| YY1 | 0.260 | 0.294 | 0.274 |

**Supplementary table 3:**
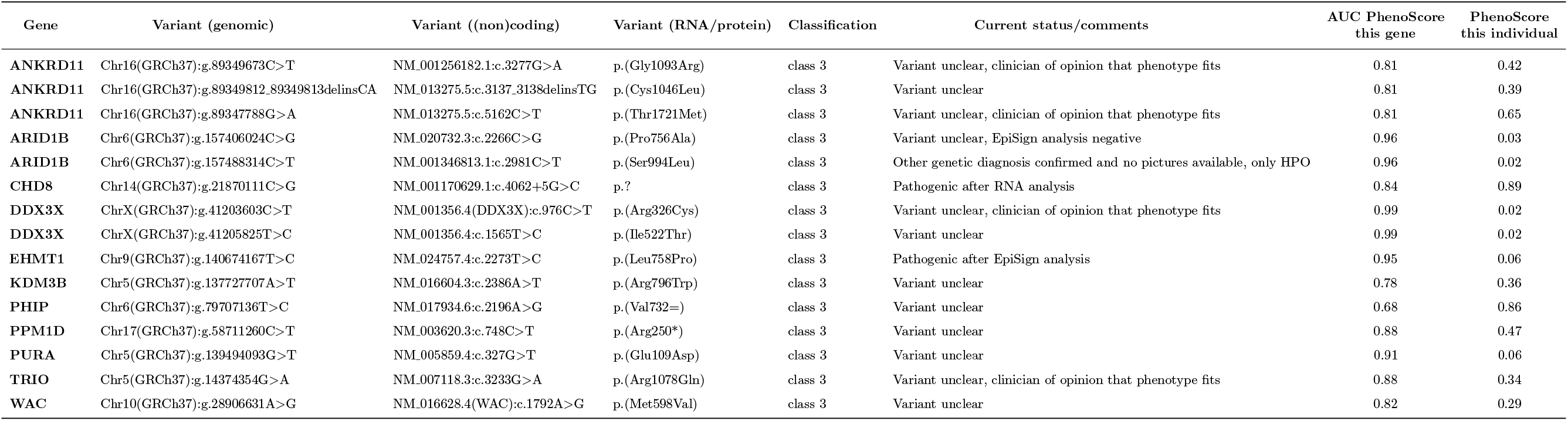
The 15 individuals with a VUS in one of the 26 included syndromes are displayed here, including the genetic information and the PhenoScore. For most, pathogenicity is still not clear at the time of writing, but for four, additional (genetic) testing has led to a change in pathogenicity class. The two *ARID1B* variants were both regarded as benign: one after methylation analysis (negative), the other variant since the individual was diagnosed with fragile X syndrome at a later stage. PhenoScore agrees with both assessments with a low prediction (0.03 and 0.02). Next to that, a splice variant in *CHD8* with a high PhenoScore of 0.89 was deemed pathogenic after RNA analysis was performed. Finally, a variant in *EHMT1* was deemed pathogenic after methylation analysis. This is the only variant in which PhenoScore disagrees with the outcome of a functional test, with a low score (0.06) – probably due to the phenotype not particularly matching.

